# Survival-Convolution Models for Predicting COVID-19 Cases and Assessing Effects of Mitigation Strategies

**DOI:** 10.1101/2020.04.16.20067306

**Authors:** Qinxia Wang, Shanghong Xie, Yuanjia Wang, Donglin Zeng

## Abstract

Countries around the globe have implemented unprecedented measures to mitigate the coronavirus disease 2019 (COVID-19) pandemic. We aim to predict COVID-19 disease course and compare effectiveness of mitigation measures across countries to inform policy decision making. We propose a robust and parsimonious survival-convolution model for predicting key statistics of COVID-19 epidemics (daily new cases). We account for transmission during a pre-symptomatic incubation period and use a time-varying effective reproduction number (*R_t_*) to reflect the temporal trend of transmission and change in response to a public health intervention. We estimate the intervention effect on reducing the infection rate and quantify uncertainty by permutation. In China and South Korea, we predicted the entire disease epidemic using only data in the early phase (two to three weeks after the outbreak). A fast rate of decline in *R_t_* was observed and adopting mitigation strategies early in the epidemic was effective in reducing the infection rate in these two countries. The lockdown in Italy did not further accelerate the speed at which the infection rate decreases. The effective reproduction number has staggered around *R_t_* = 1.0 for more than 2 weeks before decreasing to below 1.0, and the epidemic in Italy is currently under control. In the US, *R_t_* significantly decreased during a 2-week period after the declaration of national emergency, but afterwards the rate of decrease is substantially slower. If the trend continues after May 1, the first wave of COVID-19 may be controlled by July 26 (CI: July 9 to August 27). However, a loss of temporal effect on infection rate (e.g., due to relaxing mitigation measures after May 1) could lead to a long delay in controlling the epidemic (November 19 with less than 100 daily cases) and a total of more than 2 million cases.

## 1 Introduction

COVID-19 pandemic is currently a daunting global health challenge. The novel coronavirus was observed to have a long incubation period and highly infectious during this period^1-4^. The cumulative case number surpasses 4.1 million by May 10, with more than 1.3 million in the United States (US). It is imperative to study the course of the disease outbreak in countries that have controlled the outbreak (e.g., China and South Korea) and compare mitigation strategies to inform decision making in regions that are in the midst of (e.g., the US) or at the beginning of outbreak (e.g., South America).

Various infectious disease models are proposed to estimate the transmission of COVID-19^5-7^ and investigate the impact of public health interventions on mitigating the spread^8–12^. Several studies modeled the transmission by stochastic dynamical systems^5–7,10^, such as susceptible-exposed-infectious-recovered (SEIR) models^5^, extended Kalman filter^13–15^, and individual-based simulation models^8,9^. Some models did not explicitly take into account of behavioral change (e.g., social distancing) and government mitigation strategies that can have major influences on the disease course, while other work modified the infection rate as public-health-intervention-dependent^10,12^ or time-varying^7^. A recent study^11^ considered the disease incubation period and used a convolution model based on SEIR. A state-space susceptible-infectious-recovered (SIR) model with time-varying transmission rate^16^ was developed to account for interventions and quarantines.

SEIR models can incorporate mechanistic characteristics and scientific knowledge of virus transmission to provide useful estimates of its temporal dynamics, especially when individual-level epidemiological data are available through surveillance and contact tracing. However, these sophisticated models may involve a large number of parameters and assumptions about individual transmission dynamics. Thus, they may be susceptible to perturbation of parameters and prior assumptions, yielding wide prediction intervals especially when granular individual-level data are not available. In contrast to infectious disease models, alternative statistical models are proposed to predict summary statistics such as deaths and hospital demand under a nonlinear mixed effects model framework^17^, survival analysis has been introduced to model the occurrence of clinical events in infectious disease studies^18^, and a nonparametric space-time transmission model was developed to incorporate spatial and temporal information for predictions at the county level^19^. Nonparametric modelling or survival models are data-driven, so parameters may not be scientifically related to disease epidemic.

In this work, we propose a parsimonious and robust population-level survival-convolution model that is based on main characteristics of COVID-19 epidemic and observed number of confirmed cases to predict disease course and assess public health intervention effect. Our method models only key statistics (e.g., daily new cases) that reflect the disease epidemic over time with at most six parameters, so it may be more robust than models that rely on individual transmission processes or a large number of parameters and assumptions. We construct our model based on prior scientific knowledge about COVID-19, instead of posthoc observations of the trend of disease spread. Specifically, two important facts we consider include (1) SARS-CoV-2 virus has an incubation period up to 14-21 days^1^ and a patient can be highly infectious in the pre-symptomatic phase; (2) infection rate varies over time and can change significantly when government guidelines and mitigation strategies are implemented; (3) intervention effect may be time-varying.

We aim to achieve the following goals. The first goal is to fit observed data to predict daily new confirmed cases and latent pre-symptomatic cases, the peak date, and the final total number of cases. The second goal is to assess the effect of nationwide major interventions across countries (e.g., mitigation measures) under the framework of natural experiments (e.g., longitudinal pre-post quasi-experimental design^20^). Quasi-experiment approaches are often used to estimate intervention effect of a public health intervention (e.g., HPV vaccine ^21^) or a health policy where randomized controlled trials (RCTs) are not feasible. Our third goal is to project the future trend of COVID-19 for the countries (e.g., US) amid the epidemic under different assumptions of future infection rates, including the continuation of the current trend and relaxing mitigation measures.

## 2 Methods

### 2.1 Data source

We used data from a publicly available database that consolidates multiple sources of official reports (World Meters[https://www.worldometers.info/coronavirus/]). We analyzed two countries with a large number of confirmed cases in Asia (China, South Korea) and two outside (Italy, US). Since both China and South Korea are already at the end of epidemic, we used their data to test empirical prediction performance of our method. We included data in the early phase of epidemic as training set to estimate model parameters and leave the rest of the data as testing set for evaluation. For China, we used data up to two weeks post the lockdown of Wuhan city (January 23) as training (data from January 20 to February 4), and used the remaining observed data for evaluation (February 5 to May 10). Similarly, for South Korea we used data from February 15 to March 4 as training and leave the rest for evaluation (March 5 to May 10). Italy is the first European country confronted by a large outbreak and currently has passed its peak. We estimate the effect of the nation-wide lockdown in Italy (dated March 11) using 10 weeks data (February 20 to April 29). For the US, since after May 1 some mitigation measures were lifted in various states, we also included about 10 weeks data (February 21 to May 1) to assess the effect of its mitigation strategies.

### 2.2 Survival-Convolution Model

Let *t* denote the calendar time (in days) and let *N*_0_(*t*) be the number of individuals who are newly infected by COVID-19 at time *t*. Let *t_j_* denote the time when individual *j* is infected (*t_j_* = ∞ if never infected), and let *T_j_* be the duration of this individual remaining infectious to any other individual and in the transmission chain. Let *t*_0_ be the unknown calendar time when the first patient (patient zero) is infected. Therefore, at time *t*, the total number of individuals who can infect others is 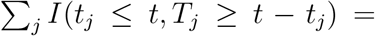 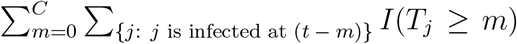, where *C* = min(*t* − *t*_0_,*C*_1_) with *C*_1_ as the maximum incubation period (i.e., 21 days for SARS-CoV-2) and *I*(*E*) denotes an indicator function with *I*(*E*) = 1 if event *E* occurs and *I*(*E*) = 0 otherwise. Since the total number of individuals who are newly infected at time (*t* − *m*) is *N*_0_(*t* − *m*), the number of individuals who remain infectious at time *t* is 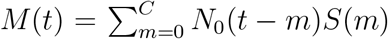, where *S*(*m*) denotes the proportion of individuals remaining infectious after *m* days of being infected, or equivalently, the survival probability at day *m* for *T_j_*. On the other hand, right after time *t*, some individuals will no longer be in the transmission chain (e.g., due to testing positive and quarantine or out of infectious period) with duration *T_j_* = (*t* − *t_j_*). The total number of these individuals is 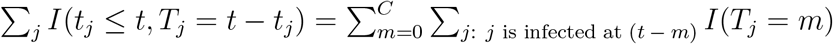, or equivalently

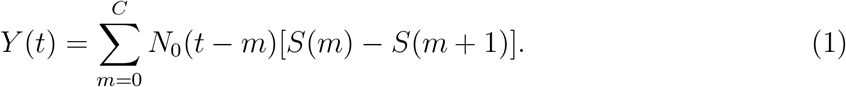

Therefore, (*M*(*t*) − *Y*(*t*)) is the number of individuals who can still infect others after time *t*. Assuming the infection rate at *t* to be *a*(*t*), then at time (*t* + 1) the number of newly infected patients is *a*(*t*)[*M*(*t*) − *Y*(*t*)], which yields

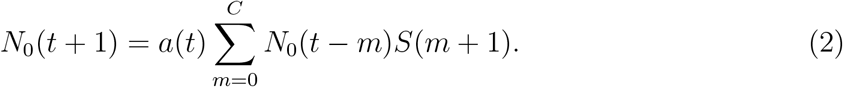

Note that *a*(*t*) is time-varying because the infection rate depends on how many close contacts an infected individual may have at time *t*, which is affected by public heath interventions (e.g., stay-at-home order, lockdown), and saturation level of the infection in the whole population. Define 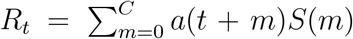, the expected number of secondary cases infected by a primary infected individual in a population at time *t* while accounting for the entire incubation period of the primary case. Thus, *R_t_* is the instantaneous time-varying effective reproduction number^22^ that measures temporal changes in the disease spread.

Models (1) and (2) provide a robust dynamic model to characterize COVID-19 epidemic. Equation (2) gives a convolution update for the new cases using the past numbers, while equation (1) gives the number of cases out of transmission chain at time *t*, and *M*(*t*) computes the number of latent pre-symptomatic cases by the end of time *t*. This model considers three important quantities to characterize COVID-19 transmission: the initial date, *t*_0_, of the first (likely undetected) case in the epidemic, the survival function of time to out of transmission, *S*(*m*), and the infection rate over calendar time, *a*(*t*).

We model infection rate *a*(*t*) as a non-negative, piece-wise linear function with knots placed at meaningful event times. The simplest model consists of a constant and a single linear function with three parameters (infection date of patient zero, intercept and slope of *a*(*t*)). When a massive public health intervention (e.g., nation-wide lockdown) is implemented at some particular date, we introduce an additional linear function afterwards with a new slope parameter. Thus, the difference in slope parameters of *a*(*t*) before and after an intervention reflects its effect on reducing the rate of change in disease transmission (i.e., “flattening the curve”). Since the intervention effect may diminish over time, we introduce another slope parameter two weeks after intervention to capture the longer-term effect. We use existing knowledge of SARS-CoV-2 virus incubation period^1^ to approximate *S*(*m*) and perform sensitivity analysis assuming different parameters. For estimation, we minimize a loss function measuring differences between model predicted and observed daily number of cases. For statistical inference, we use permutation based on standardized residuals. All mathematical details are in Supplementary Material.

### 2.3 Utility of Our Model

First, with parameters estimated from data and assuming that the future infection rate remains the same trend, we can use models (1) and (2) to predict future daily new cases, the peak time, expected number of cases at the peak, when *R_t_* will be reduced to below 1.0, and when the epidemic will be controlled (the number of daily new cases below a threshold or decreases to zero). Furthermore, our model provides the number of latent cases cumulative over the incubation period at each future date, which can be useful to anticipate challenges and allocate resources effectively.

Second, we can estimate the effects of mitigation strategies, leveraging the nature of quasi-experiments where subjects receive different interventions before and after the initiation of the intervention. The longitudinal pre-post intervention design allows valid inferences assuming that pre-intervention disease trend would have continued had the intervention not taken place and local randomization holds (whether a subject falls immediately before or after the initiation date of an intervention may be considered as random, and thus the “intervention assignment” may be considered to be random). Applying this design, the intervention effects will be estimated as the difference in the rate of change of the infection rate function before and after an intervention takes place.

Third, we study the impact of an intervention (e.g., lifting mitigation measures) that changes the epidemic at a future date. Using permutations, we obtain the joint distribution of the parameter estimators and construct confidence intervals (CI) for the projected case numbers and interventions effects.

## 3 Results

For China, the infection rate *a*(*t*) is a single linear function (estimates in Table 1). The first community infection was estimated to occur on January 3, 17 days before the first reported case (Table 1). Figure 1A shows that the model captures the peak date of new cases, the epidemic end date, and the prediction interval contains the majority of observed number of cases except one outlier (due to a change of diagnostic criteria). The reproduction number *R_t_* decreases quickly from 3.34 to below 1.0 in 14 days (Figure 2A). We only used data up to February 4 to estimate our model. The observed total number of cases by May 10 is 82,901, which is inside the 95% CI of the estimated total number of cases (58,415; 95% CI: (42,516, 133,083)). There are two outlier days (February 12, 13) with a total of 19,198 cases reported in the testing set. Excluding two outliers, the observed number of cases 62,356.

**Table 1:**
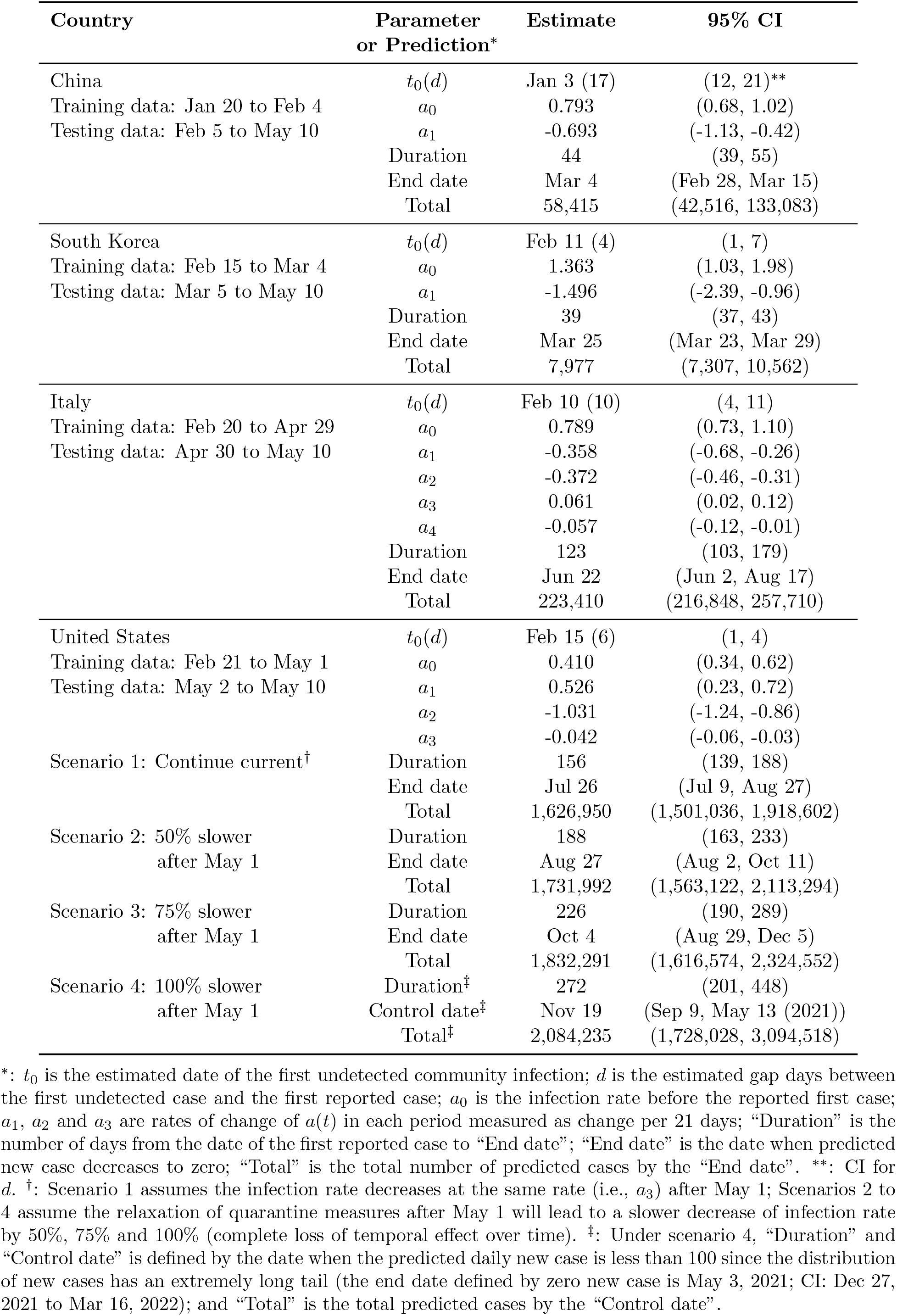
Model Estimated Parameters in Each Country

| Country | Parameter<br>or Prediction* | Estimate | 95% CI |
| --- | --- | --- | --- |
| China | $t_0(d)$ | Jan 3 (17) | (12, 21)** |
| Training data: Jan 20 to Feb 4 | $a_0$ | 0.793 | (0.68, 1.02) |
| Testing data: Feb 5 to May 10 | $a_1$ | -0.693 | (-1.13, -0.42) |
|  | Duration | 44 | (39, 55) |
|  | End date | Mar 4 | (Feb 28, Mar 15) |
|  | Total | 58,415 | (42,516, 133,083) |
| South Korea | $t_0(d)$ | Feb 11 (4) | (1, 7) |
| Training data: Feb 15 to Mar 4 | $a_0$ | 1.363 | (1.03, 1.98) |
| Testing data: Mar 5 to May 10 | $a_1$ | -1.496 | (-2.39, -0.96) |
|  | Duration | 39 | (37, 43) |
|  | End date | Mar 25 | (Mar 23, Mar 29) |
|  | Total | 7,977 | (7,307, 10,562) |
| Italy | $t_0(d)$ | Feb 10 (10) | (4, 11) |
| Training data: Feb 20 to Apr 29 | $a_0$ | 0.789 | (0.73, 1.10) |
| Testing data: Apr 30 to May 10 | $a_1$ | -0.358 | (-0.68, -0.26) |
| | $a_2$ | -0.372 | (-0.46, -0.31) |
| | $a_3$ | 0.061 | (0.02, 0.12) |
| | $a_4$ | -0.057 | (-0.12, -0.01) |
|  | Duration | 123 | (103, 179) |
|  | End date | Jun 22 | (Jun 2, Aug 17) |
|  | Total | 223,410 | (216,848, 257,710) |
| United States | $t_0(d)$ | Feb 15 (6) | (1, 4) |
| Training data: Feb 21 to May 1 | $a_0$ | 0.410 | (0.34, 0.62) |
| Testing data: May 2 to May 10 | $a_1$ | 0.526 | (0.23, 0.72) |
| | $a_2$ | -1.031 | (-1.24, -0.86) |
| | $a_3$ | -0.042 | (-0.06, -0.03) |
| Scenario 1: Continue current<sup>†</sup> | Duration | 156 | (139, 188) |
|  | End date | Jul 26 | (Jul 9, Aug 27) |
|  | Total | 1,626,950 | (1,501,036, 1,918,602) |
| Scenario 2: 50% slower<br>after May 1 | Duration | 188 | (163, 233) |
|  | End date | Aug 27 | (Aug 2, Oct 11) |
|  | Total | 1,731,992 | (1,563,122, 2,113,294) |
| Scenario 3: 75% slower<br>after May 1 | Duration | 226 | (190, 289) |
|  | End date | Oct 4 | (Aug 29, Dec 5) |
|  | Total | 1,832,291 | (1,616,574, 2,324,552) |
| Scenario 4: 100% slower<br>after May 1 | Duration<sup>‡</sup> | 272 | (201, 448) |
|  | Control date<sup>‡</sup> | Nov 19 | (Sep 9, May 13 (2021)) |
|  | Total<sup>‡</sup> | 2,084,235 | (1,728,028, 3,094,518) |

\*: $t_0$ is the estimated date of the first undetected community infection; $d$ is the estimated gap days between the first undetected case and the first reported case; $a_0$ is the infection rate before the reported first case; $a_1$ , $a_2$ and $a_3$ are rates of change of $a(t)$ in each period measured as change per 21 days; “Duration” is the number of days from the date of the first reported case to “End date”; “End date” is the date when predicted new case decreases to zero; “Total” is the total number of predicted cases by the “End date”. \*\*: CI for $d$ . †: Scenario 1 assumes the infection rate decreases at the same rate (i.e., $a_3$ ) after May 1; Scenarios 2 to 4 assume the relaxation of quarantine measures after May 1 will lead to a slower decrease of infection rate by 50%, 75% and 100% (complete loss of temporal effect over time). ‡: Under scenario 4, “Duration” and “Control date” is defined by the date when the predicted daily new case is less than 100 since the distribution of new cases has an extremely long tail (the end date defined by zero new case is May 3, 2021; CI: Dec 27, 2021 to Mar 16, 2022); and “Total” is the total predicted cases by the “Control date”.

**Figure 1:**
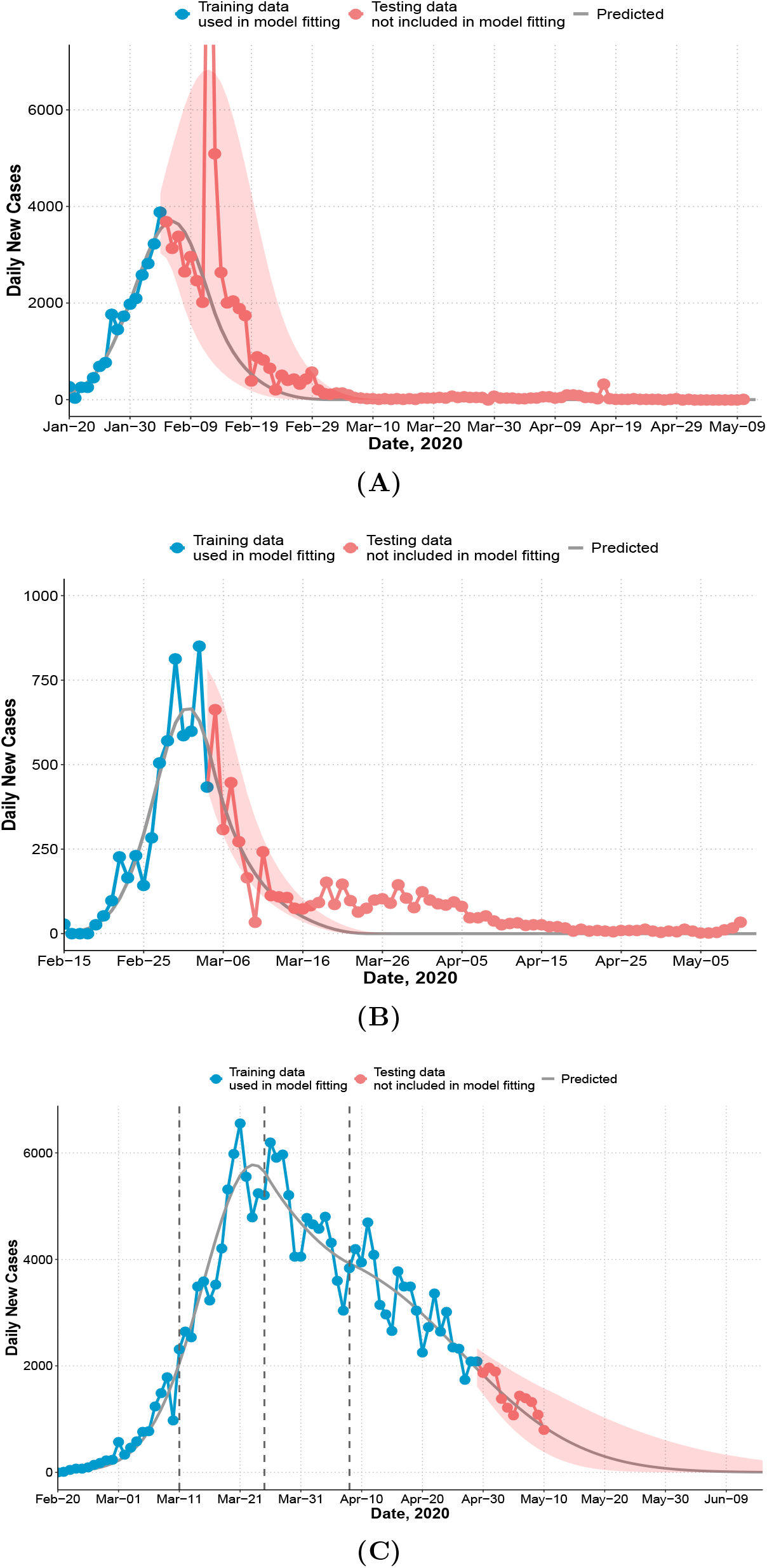
Observed and predicted daily new cases and 95% prediction interval (shaded). (**A**) China. Training data: January 20 to February 4; testing data: February 5 to May 10. 14,108 cases were reported on February 12 and not shown on figure. The recent cases since April are imported cases. (**B**) South Korea. Training data: February 15 to March 4; testing data: March 5 to May 10. (**C**) Italy. First dashed line indicates the nation-wide lockdown (March 11). Second and third dashed line indicates two or four weeks after. Training data: February 20 to April 29 (7 weeks after the lockdown); testing data: April 30 to May 10.

For South Korea, Figure 1B shows that the model captures the general trend of the epidemic except at the tail area (after March 15) where some small and enduring outbreak is observed. The effective reproduction number decreases dramatically from 5.37 at the beginning of the outbreak to below 1.0 in 14 days (Figure 2B). The predicted number of new cases at the peak is 665 and the total number of predicted cases at the peak time is close to the observed total (4,300 vs 4,335). The predicted total number by March 15 is 7,816 and the observed total is 8,162.

**Figure 2:**
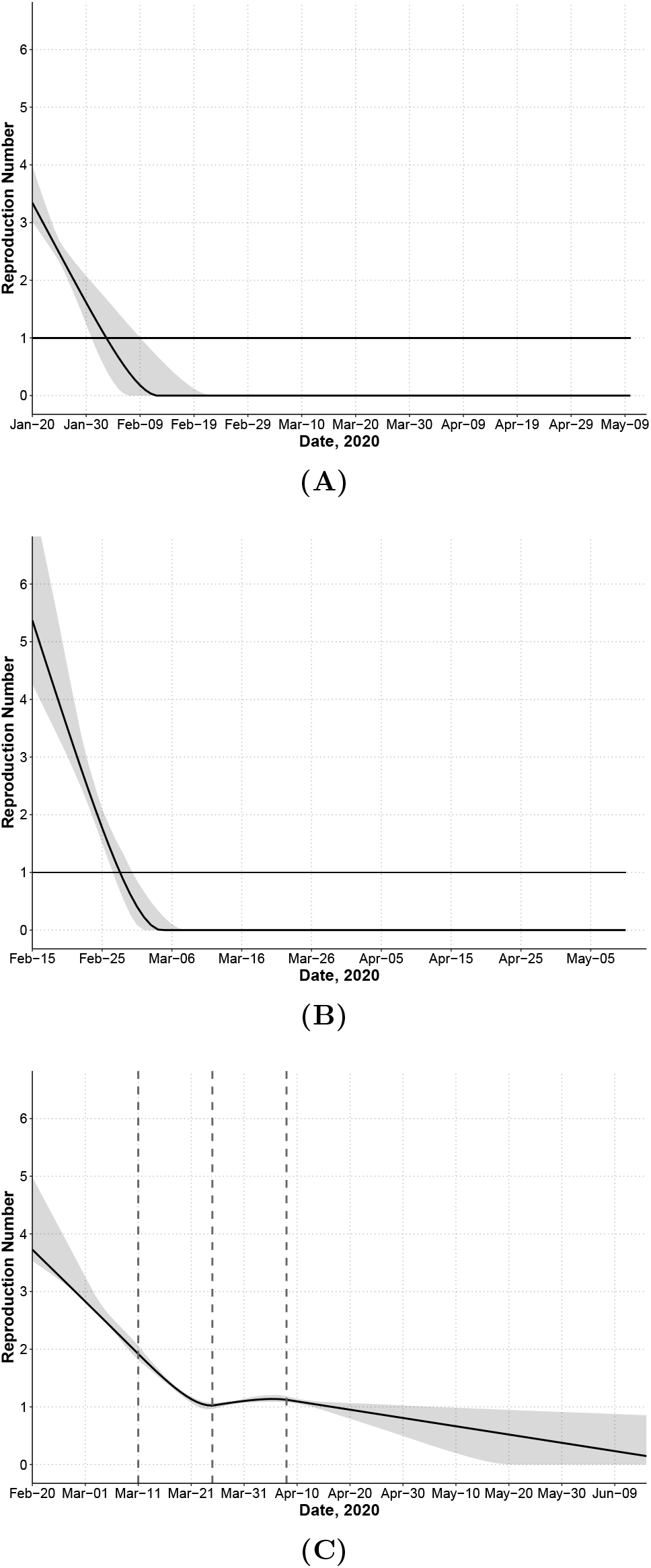
Effective reproduction number *R_t_* for each country computed as the average number of secondary infections generated by a primary case at time *t* accounting for the incubation period of the primary case. Dashed lines indicate knots for infection rate *a*(*t*). (**A**) China. (**B**) South Korea. (**C**) Italy.

For Italy, we model *a*(*t*) as a four-piece linear function to account for the change in mitigation strategies with a knot placed at the lockdown (March 11), and two additional knots at 2-week intervals (March 25, April 8) to account for time-varying intervention effect. Difference on the rate of change before and after the first knot measures the immediate effect of lockdown on reducing the infection rate. Change before and after the second and third knot measures whether the lockdown effect can be maintained in longer term. The rate of change in *R_t_* is not significantly different before and two weeks after the lockdown (Figure 2C). The reproduction number decreased from 3.73 at the beginning to 1.02 two weeks post-lockdown. However, starting from the third week post-lockdown (March 26), *R_t_* stops decreasing and remains close to 1.0 until April 16. The slope of *a*(*t*) (infection rate) increases by 116% to a slightly positive value after March 26 (Table 1, comparing *a*_2_ and *a*_3_ for Italy). This is consistent with a relatively flat trend of observed daily new cases during this period (Figure 1C). The estimated total by May 10 is 216,300 (95%CI: (214,863, 228,406)) and close to the observed total (219,070). Recent daily cases in the testing set also closely follow our predicted trend (Figure 1C).

In the US, we fit a three-piece model for *a*(*t*) with a knot on March 13 (the declaration of national emergency) and an additional knot two weeks after (March 27). The predicted peak date is May 3 (Figure 3A) with a total number of 1,176,915 cases by May 3, which is close to the observed total (1,188,122). *R_t_* increases during the early phase but decreases sharply after the declaration of national emergency (Figure 3B) up to two weeks after. During the next period (March 28 to April 10), *R_t_* decreases at a much slower rate. If this trend continues, the end of epidemic date is predicted to be July 26 (scenario 1, Figure 3A), and the predicted total over the entire epidemic will be 1,626,950 (CI: (1,501,036, 1,918,602), Table 1). However, since states in the US are gradually lifting mitigation measures after May 1, the trend of infection rate may change. We predicted epidemic control date assuming *a*(*t*) decreases slower after May 1 by 50% (scenario 2), 75% (scenario 3), and 100% (scenario 4) in Table 1. Under scenario 4 where the temporal effect of mitigation measures is completely lost (i.e., *a*(*t*) is a constant over time), the projected total number of cases will be more than 2 million, and the epidemic cannot be controlled until November 19 (with less than 100 daily cases, Table 1). Assuming a case fatality rate of 6% as observed by May 10, the total number of deaths would be around 120,000.

**Figure 3:**
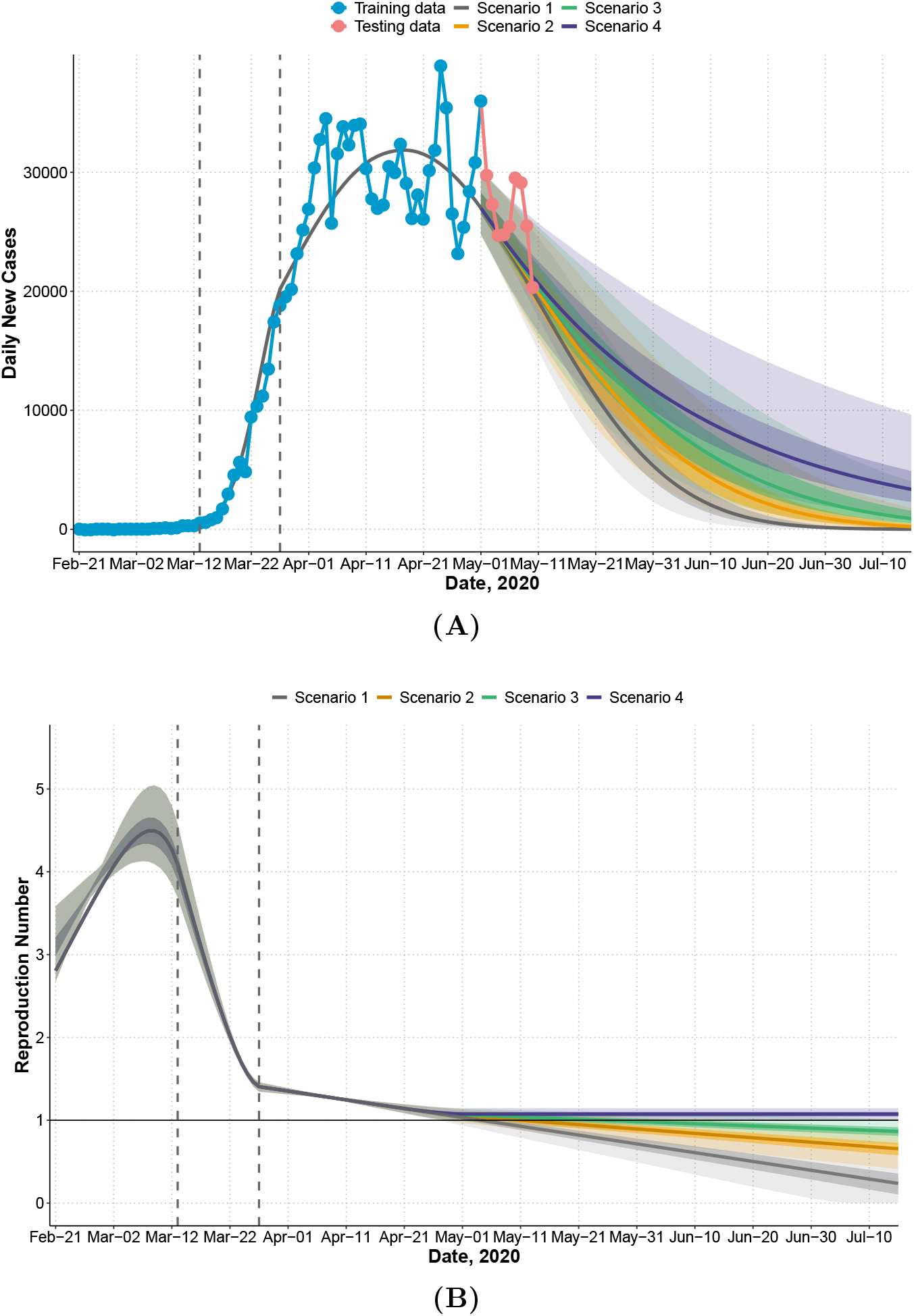
United States: observed and predicted daily new cases, 95% prediction intervals (lighter shaded) and 50% prediction intervals (darker shaded) under four scenarios that assume relaxation of mitigation measures occurs after May 1. Scenario 1: infection rate *a*(*t*) follows the same trend after May 1 as observed between March 27 and May 1. Scenario 2: rate of decrease of *a*(*t*) slows by 50% after May 1. Scenario 3: rate of decrease of *a*(*t*) slows by 75% after May 1. Scenario 4: rate of decrease of *a*(*t*) slows by 100% after May 1 (complete loss of temporal decreasing effect). First dashed line indicates the declaration of national emergency (March 13). Second dashed line indicates two weeks after (March 27). Training data: February 21 to May 1 (7 weeks after declaring national emergency); testing data: May 2 to May 10. (**A**) Observed and predicted daily new cases. (**B**) Effective reproduction number *R_t_*.

We show the estimated number of latent cases present on each day (i.e., including pre-symptomatic patients infected *k* days before but have not shown symptoms) in Supplementary Material (Figure S1). For all countries, there were a large number of latent cases around the peak time. We performed a sensitivity analysis using different distributions of *S*(*m*) assuming a delay in reporting confirmed cases. The results show that predicted daily new cases were similar under different parameters of *S*(*m*) for both US and Italy (Supplementary Material Figures S2 and S3), demonstrating robustness of our method to the assumptions of *S*(*m*).

## 4 Discussion

In this study, we propose a parsimonious and robust survival convolution model to predict daily new cases of the COVID-19 outbreak and use a natural quasi-experimental design to estimate the effects of mitigation measures. Our model accounts for major characteristics of COVID-19 (long incubation period and highly contagious during incubation) with a small number of parameters (up to six) and assumptions, directly targets prediction accuracy, and provides measures of uncertainty and inference based on permuting the residuals. We allow the infection rate to depend on time and modify the basic reproduction number *R*_0_ as a time-dependent measure *R_t_* to estimate change in disease transmission over time. Thus, *R_t_* corrects for the naturally impact of time on the disease spread. Our estimated reproduction number at the beginning of the epidemic ranges from 2.81 to 5.37, which is consistent with R_0_ reported in other studies^23^ (range from 1.40 to 6.49, with a median of 2.79). For predicting daily new cases, our analyses suggest that the model estimated from early periods of outbreak can be used to predict the entire epidemic if the disease infection rate dynamic does not change dramatically over the disease course (e.g., about two weeks data is sufficient for China and fits the general trend of South Korea).

Comparing the effective reproduction numbers across countries, *R_t_* decreased much more rapidly in South Korea and China than Italy (Figure 2). In South Korea, the effective reproduction number had been reduced from 5.37 to under 1.0 in a mere 13 days and the total number of cases is low. The starting reproduction number in South Korea was high possibly due to many cases linked to patient 31 and outbreaks at church gatherings. Similarly for China, the reproduction number reduced to below 1.0 in 14 days. Italy’s *R_t_* decreased until almost reaching 1.0 on March 25, but remained around 1.0 for 3 weeks. The US followed a fast decreasing trend during a two-week period after declaring national emergency (*a*_2_ = −1.031), which is faster than the first two weeks in China (*a*_1_ = −0.693), but its R_t_ decreased at a much slower rate (*a*_3_ = −0.042) afterwards and was below 1.0 on May 5.

Comparing mitigation strategies across countries, the fast decline in R in China suggests that the initial mitigation measures put forth on January 23 (lockdown of Wuhan city, traffic suspension, home quarantine) were successful in controlling the transmission speed of COVID-19. Additional mitigation measures were in place after February 2 (centralized quarantine and treatment), but did not seem to have significantly changed the disease course. In fact, our model assuming the same infection rate trajectory after February 2 fits all observed data up to May 10. A recent analysis of Wuhan’s data^24,25^ arrived at a similar conclusion, and their estimated *R_t_* closely matches with our estimates. However, their analyses were based on self-reported symptom onset and other additional surveillance data, where we used only widely available official reports of confirmed cases. Another mechanistic^26^ study confirmed the effectiveness of early containment strategies in Wuhan.

South Korea did not impose a nation-wide lockdown or closure of businesses, but at the very early stage (when many cases linked to patient 31 were reported on February 20) conducted extensive broad-based testing and detection (drive through tests started on February 26), rigorous contact tracing, isolation of cases, and mobile phone tracking. Our results suggest that South Korea’s early mitigation measures were also effective.

Italy’s initial mitigation strategies in the most affected areas reduced *R_t_* from 3.73 to 1.92 in 20 days. To estimate the intervention effect of the nation-wide lockdown as in a natural experiment, we require local randomization and the continuity assumption. The former requires that characteristics of subjects who are infected right before or after the lockdown are similar. Since in a very short time period, whether a person is infected at time *t* or *t* + 1 is likely to be random, the local randomization assumption is likely to be valid. Continuity assumption refers to that the infection rate before the lockdown would continue to capture the trend afterwards had the intervention not been implemented. Under this assumption, the lockdown in Italy is not effective to further reduce the transmission speed (slopes of *a*(*t*) are similar before and after lockdown on March 11). There were 10,149 cases reported in Italy as of March 10, suggesting that the lockdown was placed after the wide community spread had already occurred. Nevertheless, it is possible that without the lockdown the infection rate would have had increased, i.e., the lockdown enhanced and maintained the effect of quarantine for two weeks. In fact, after two weeks of lockdown, we observe a loss of temporal effect so that *R_t_* has remained around 1.0 for about 2-3 weeks before it starts to decrease again.

For the US, *R_t_* ranges between 2.81 and 4.50 before the declaration of national emergency on March 13, but *R_t_* declines rapidly over a two-week period after March 13. Although the disease trend and mitigation strategies vary across states in the US, since the declaration of national emergency, many states have implemented social distancing and ban of large gathering. The large difference before and after March 13 is likely due to states with large numbers of cases that implemented state-wide mitigation measures (e.g., New York, New Jersey). Our model predicted a continued decrease in *R_t_* from March 27 to May 1 but at a much slower rate (95.9% slower; Table 1, comparing *a_2_* and *a_3_* for the US). If the trend continues after May 1, the first wave of epidemic will be controlled by July 26 (CI: July 9, August 27). However, after May 1 many states enter a re-opening phase. If the guidelines on quarantine measures are relaxed so that the effect of quarantine cannot be maintained, the control date can be delayed by 32 days (50% slower decrease in the infection rate) or 70 days (75% slower). If the temporal effect of quarantine measures is completely lost, the predicted total number of cases is more than 2 million, with a long delay in controlling the epidemic (less than 100 cases by November 19, and no new case by May, 2021).

Other studies reported transmission between asymptomatic individuals^6^, which is not accounted for here. However, asymptomatic individuals can only be identified and confirmed by serological tests which are not widely available. When there is a delay in reporting some symptomatic patients, the daily reported cases are a mixture of new symptomatic cases and patients presenting after having had symptoms for a few days. In this case, the average number of days to testing positive may be higher than the virus incubation period of 5.2 days. However, as shown in our sensitivity analysis, the prediction of daily reported cases was not affected by using a larger mean value for *S*(*m*), demonstrating robustness of the model. Our model does not consider subject-specific covariates and focuses on predicting population-level quantities. Neither have we considered borrowing information from multiple countries or state-level analysis for the US, which are worthy of study in a mixed effects model framework. We do not consider prediction of daily new deaths or hospitalizations. These data can be included to enhance the prediction of new cases by linking the distribution of time to COVID symptom onsets, hospitalization, or death. Lastly, we can consider a broader class of models for infection rate *a*(*t*) to allow discontinuity in both intercepts and slopes before and after an intervention under a regression discontinuity design^21,27^.

Despite these limitations, our study offers several implications. Implementing mitigation measures earlier in the disease epidemic reduces the disease transmission rate at a faster speed (South Korea, China). Thus for regions at the early stage of disease epidemic, mitigation measures should be introduced early. Nation-wide lockdown may not further reduce the speed of *R_t_* reduction compared to regional quarantine measures as seen in Italy. In countries where disease transmissions have slowed down, lifting of quarantine measures may lead to a persistent infection rate delaying control of epidemic and thus should be implemented with caution and close monitoring.

## Data Availability

All our data and optimization codes are publicly available at https://github.com/COVID19BIOSTAT/covid19_prediction

https://github.com/COVID19BIOSTAT/covid19_prediction

## Data sharing

All data and optimization codes are publicly available at [https://github.com/COVID19BIOSTAT].

## Acknowledgements

The authors are funded in part by the US NIH grants NS073671, GM124104, and MH117458.

